# Women’s intentions and motivations towards health behaviour change before pregnancy: a cross-sectional survey of pregnant women in Australia

**DOI:** 10.64898/2026.06.11.26355496

**Authors:** Amie Steel, Danielle Schoenaker, Erica McIntyre, Kris Rogers, Jennifer Hall, Jon Adams

## Abstract

**Introduction:** The preconception period (i.e., the weeks and months before pregnancy) is a critical window during which parental health behaviours can influence pregnancy outcomes and the child’s long-term health. Modifiable factors such as nutrition, physical activity, substance use, and environmental exposures play a key role, yet women’s ability to adopt and sustain healthy behaviours is shaped by complex psychological, social and environmental influences. This study applies the Theory of Planned Behaviour to identify the beliefs underpinning women’s preconception behaviours, with the aim of informing support for effective and sustained health behaviour change.

**Methods:** An Australian national retrospective cross-sectional survey of pregnant women (18-49 years), recruited through social media platforms. The 92-item survey captured respondent socio-demographics, pregnancy status and health conditions, health behaviours, and beliefs regarding preconception health behaviours. Respondents’ level of pregnancy planning was categorised using the London Measure of Unplanned Pregnancy (LMUP). Items regarding preconception beliefs were structured in accordance with the Theory of Planned Behaviour, with a focus on regular exercise, healthy diet, and alcohol avoidance. These beliefs variables were analysed using structured equation modelling to identify paths between latent variables and the items used to estimate each concept.

**Results:** The study was completed by 430 pregnant women of whom 72.7% had a planned pregnancy. Most had a partner, were university educated and in good health. Structural equation modelling showed intention strongly predicted exercise (β=0.65), healthy diet (β=0.54) and alcohol avoidance (β=0.64). Perceived control and partner norms influenced intentions, whereas health professional norms had limited effect. Positive beliefs were associated with folate supplement use and smoking cessation.

**Conclusion:** These findings highlight intention as a key driver of preconception health behaviours, with perceived control and partner influences playing a more significant role than individual beliefs or health professional input. Effective interventions should therefore address structural barriers and actively involve partners, while respecting women’s autonomy. Overall, couples-focused, multi-level strategies are likely essential to support meaningful and sustained preconception health behaviour change.

## Introduction

The weeks and months before pregnancy – referred to as the ‘preconception period’ – is a critical time in which a couple’s health status and behaviours can impact the pregnancy outcome and the health of the child at birth and later in life (Stephenson et al., 2018). A range of modifiable risk factors in both parents can impact on pregnancy and birth outcomes, including body composition, substance use (alcohol, smoking, illicit drugs), physical activity, nutrient intake, and environmental exposures (Carter et al., 2023; Caut et al., 2022). Adverse outcomes associated with these exposures can impact the health of the mother during pregnancy or developing embryo as well as have direct effects on the offspring (Caut et al., 2022). For this reason, couples planning a pregnancy should be supported to improve health behaviours and ameliorate risk exposures as early as possible before conception.

The success with which women can implement and sustain health behaviour change during the preconception period, however, can be influenced by a range of internal (e.g., psychological, biological) and external (e.g., environmental, social, cultural) factors (Kandel et al., 2021; Rockliffe et al., 2021; Steel et al., 2025). The complex interplay between these factors means that meaningful support for women to improve their preconception health relies on interventions across health policy, health promotion and health services (Steel et al., 2025). They also may rely on influencing the attitudes and behaviours of women and their social networks, including their reproductive partner, to ensure they are fully supported to make appropriate changes while still retaining bodily autonomy (DeBruin & Marshall, 2019; Rockliffe et al., 2021). Yet many preconception health behaviour interventions rely on the involvement of a health professional and target women alone rather than including both reproductive partners (Hemsing et al., 2017; Withanage et al., 2022). To better facilitate women’s preconception health behaviour change, the relationship between the different factors which may influence their behaviour decisions and ability to enact them need more clarity.

## Material and methods

### Aim

This study aimed to explore the relationships between women’s beliefs, attitudes or intentions and their preconception health behaviours, using the theory of planned behaviour (Ajzen, 1991).

### Ethical clearance

The study was determined to meet the National Health and Medical Research Council’s National Ethics Statement by the Medical Research Ethics Committee of the University of Technology Sydney (#ETHI21-6461).

### Study design and participants

The study employed a cross-sectional observational survey design, using an online questionnaire. Participants were eligible to participate if there were a pregnant female currently living in Australia. They were recruited directly through social media advertisements on Facebook, Instagram and X (formerly ‘Twitter’) between December 2020 and October 2021. Boosted social media advertisements targeted Meta account holders (Facebook and Instagram) listed as female between the ages of 18 and 49 years old. A random prize draw of $100 was available to individuals who complete the survey. All data were collected using an online questionnaire constructed using the Qualtrics™ online survey platform (Qualtrics, 2025). Interested parties accessed a screening instrument via a link on the social media advert. Participants who met the eligibility criteria were sent an email containing the link for their survey and a second survey link which they were asked to forward to their reproductive partner by email. No a priori power calculation was performed. Participants were presented with a participant information sheet and consented to study participation prior to accessing the survey.

### Theoretical framework

This analysis employs the Theory of Planned Behaviour (TPB) to explore the drivers of women’s preconception health behaviours (Ajzen, 1991). The TPB postulates that an individual’s beliefs about a behaviour (i.e. their beliefs about the value or impact of the behaviour), their perceptions of norms (both personal [i.e. ‘what I think people like me do’] and subjective [i.e. ‘what I think others perceive people like me do’]), and perceptions of control (i.e. their views on their ability to influence their own behaviour), all have the potential to influence their intention to engage in that behaviour, and ultimately the behaviour itself.

### Survey instrument

The 92-item survey instrument was created based on questionnaires initially developed and utilised in the United Kingdom (Stephenson et al., 2014). The survey instrument was pilot tested for face and functional validity by three individuals who met the eligibility criteria of the target sample population. Minor adjustments to wording and survey flow were made based on their feedback. The instrument gathered information across five domains: 1) participant demographics (16 items); 2) description of current pregnancy (15 items); 3) preconception and pregnancy health behaviours, beliefs and intentions (38 items); 4) preconception and pregnancy health information and advice (13 items); and 5) health history and general health status (10 items). Domain 3 provided the critical data used for this analysis

#### Preconception and pregnancy health behaviours, beliefs and intentions

Domain 3 incorporated items from the validated Australian version of the London Measure of Unplanned Pregnancy (LMUP) (Lang et al., 2019). Behaviour variables were based on participants recent preconception behaviours and included (1) healthy eating, (2) exercising, (3) alcohol use, (4) maintained a ‘normal’ BMI one month before pregnancy, (5) took a folate supplement before pregnancy, (6) current self-reported health is ‘excellent’ or ‘good’, (7) previous smoker did not smoke 3 months before pregnancy.

This also included additional items about participant preconception health behaviours linked more directly to the timeline (i.e. 6 months prior to pregnancy) used when measuring participants’ beliefs, intentions, control, norms and subjective norms through the statement “I [behaviour] for at least 6 months before becoming pregnant”. Beliefs were then measured using a statement adapted for each behaviour from the stem statement “my doing [behaviour] during the 6 months before becoming pregnant is..” with three 7-point scale variations (0 is most affirmative and 7 is least affirmative) for each behaviour: important/not important, pleasant/unpleasant, good/bad. Intentions were measured using two statements against a 7-point scale of likelihood (extremely likely/extremely unlikely): (1) “I intend to [behaviour] for at least 6 months before becoming pregnant”, and (2) “I will make an effort to [behaviour] for at least 6 months before becoming pregnant”. Control was measured using a similar 7-point scale of likelihood using two statements: (1) “I am confident I can [behaviour] for at least 6 months before becoming pregnant”, and (2) “[Behaviour] for 6 months before becoming pregnant is up to me”. Norms were measured for personal norms (i.e. “most people like me…”) and subjective norms across different groups including reproductive partners, older female relatives, older male relatives, family and friends and health care provider.

The statements used to measure women’s beliefs, attitudes and perceptions with regards to preconception health behaviours were developed in line with the TPB (Ajzen, 1991). This theory was chosen as an established approach to understanding the relationships between individual beliefs, intentions and behaviours, particularly for health behaviours included in this analysis (Conner et al., 2001; Cooke et al., 2016; Hobbs et al., 2013; McDermott et al., 2015; Schifter & Ajzen, 1985). While these behaviours occurred in the past, they were chosen for the analysis as behaviours can change based on pregnancy outcomes and as such, these were the most proximal behaviours relevant to the study question.

The analysis in this paper uses selected survey items relevant to the research question, focusing on participant demographics, and preconception health behaviours, beliefs and attitudes.

### Statistical analysis

The study view rate was determined by the number of individuals who accessed the screening instrument and provided their email address. The survey participation percentage was calculated based on the number of people who completed the first survey item directly related to preconception health behaviour (“How long before becoming pregnant were you thinking about having a baby?”), divided by the individuals who completed the screening instrument and provided their email address. The completion rate was calculated as the number of participants completing the last survey item divided by the number who participated in the study. Participants who provided a response to the first survey item related preconception health beliefs were included in the analysis for this study. Participants without data for the items about preconception health beliefs were excluded from the analysis (n=202). Demographic differences among individuals with data included in this analysis, compared with those excluded due to missing data are presented in Appendix A (see Table A.1). Cramer’s V analysis identified moderate associations (Akoglu, 2018) between groups by highest level of education (p=0.003; V=0.1491) and holding a health care card (p=0.003; V=0.1194).

LMUP scores were calculated according to the instrument’s guidelines (Barrett, ND). Due to a technical issue with the online survey design, the final LMUP item measuring specific preconception behaviours, which should have allowed participants to select multiple options, only permitted one selection. Consequently, participants could only receive a maximum score of ‘1’ for this item, instead of the usual maximum of ‘2’. Missing data for LMUP items were managed following the LMUP scoring guide, excluding participants who responded to fewer than three items from the analysis. A new variable was created to categorise respondents’ pregnancies based on their LMUP score: ‘Planned’ (10 or more), ‘Ambivalent’ (between 4 and 9), or ‘Unplanned’ (3 or less).

Initial descriptive analysis reported continuous variables as mean (with standard deviation) or medians (with interquartile ranges), and categorical variables as frequencies and percentages.

Structural equation modelling (SEM) was employed to examine relationships among TPB constructs for alcohol use, regular exercise, and healthy eating, with latent variables specified using survey items. All SEM analyses were conducted in R (version 4.3.1) using the lavaan package (Rosseel, 2012). Model fit was evaluated using the Comparative Fit Index (CFI), Tucker–Lewis Index (TLI), Root Mean Square Error of Approximation (RMSEA), and Standardised Root Mean Square Residual (SRMR). When model convergence or satisfactory fit could not be achieved, item parcelling was implemented (16).

If acceptable fit remained unattained, TPB constructs were instead modelled using single observed indicators within a path analysis framework. Selection of indicators for the path model was independently reviewed and justified by two members of the research team, resulting in one representative variable per construct. Internal reliability analyses, using Cronbach’s alpha test of covariance between items in each scale (for scales with three or more items) or Pearson’s correlation coefficient (for scales with two items) tested consistency within the constructs. These consistency tests were used to inform the decision about selecting one item within a construct as a proxy for the whole construct for the SEM and path analyses. Regression pathways were specified to assess associations among beliefs, subjective norms, perceived behavioural control, and intention, as well as between intention and behaviour. Missing data were addressed using full information maximum likelihood estimation. Standardised path estimates are reported.

The likelihood that women’s beliefs regarding additional preconception health behaviours (i.e., maintaining a healthy weight, increasing folate intake, taking care of their health, and not smoking) predicted related health behaviours or health status (i.e. having a BMI classification of ‘normal weight’, taking a folate supplement, having ‘excellent’ or ‘good’ health, previous smokers not smoking) was estimated using logistic regression. These regressions were further adjusted for participant age, relationship status, education level, financial manageability, employment status, and cohabitation circumstances.

## Results

The survey was attempted by 908 women with 430 (47.3%) completing the survey items relevant to this analysis. Included respondents had a mean age of 30.0 years (SD 4.6) and were primarily married (55.8%) or in a de facto relationship (39.1%) (see Table 1). Most participants reported their general health status as ‘good’ (65.5%), while 32.3% reported having a longstanding illness, disability or infirmity (32.3%) (see Table 2). The most common BMI classification for participants was normal weight (44.6%) and pre-obesity (28.1%). According to the LMUP, 72.7% of women had a ‘planned’ pregnancy, and 66.8% of participants used some form of preconception folate supplement in the three months before pregnancy. Other common health behaviours before pregnancy included visiting the dentist (47.0%), checking immunisation status (37.4%), and exercise (76.5%) (see Table 3). Among the 31.6% of participants who had ever smoked, 61.8% reported not smoking in the three months prior to conception. Almost all participants reported having ever consumed alcohol (94.0%) and of those, 80.5% reported drinking alcohol in the three months before pregnancy.

**Table 1.** Participant demographics (n=430)

| **Characteristics** | **Mean (SD; min, max)** |
| --- | --- |
| Age (n=427) | 30.0 (4.6; 18, 44) |
| Sex (n=428) | **N (%)** |
| Female | 427 (99.8) |
| Male | 1 (0.2) |
| Relationship status (n=430) |  |
| Married | 240 (55.8) |
| De facto | 168 (39.1) |
| Other | 22 (5.1) |
| Highest level of education (n=430) |  |
| Year 10 or less | 11 (2.6) |
| Year 12 or equivalent | 31 (7.2) |
| Apprenticeship, certificate or diploma | 115 (26.7) |
| University qualification | 273 (63.5) |
| Financial manageability (n=430) |  |
| It is impossible/It is difficult all the time | 25 (5.8) |
| It is difficult some of the time | 95 (22.1) |
| It is not too bad | 198 (46.1) |
| It is easy | 112 (26.1) |
| Employment status (n=430) |  |
| Full time work (35 hours or more per week) | 227 (52.8) |
| Part time work (less than 35 hours per week) | 108 (25.1) |
| Casual or temporary work (irregular hours) | 37 (8.6) |
| Currently looking for work | 8 (1.9) |
| Not currently in the paid workforce and not looking for work | 50 (11.6) |
| Private health insurance cover (n=430) |  |
| No | 179 (41.6) |
| Yes, but no obstetrics cover | 138 (32.1) |
| Yes, with obstetrics cover | 113 (26.3) |
| Health Care Card* (n=430) | 87 (20.2) |
| Aboriginal or Torres Strait Islander status |  |
| No | 401 (93.3) |
| Yes | 24 (5.6) |
| I would rather not say | 5 (1.2) |
| English as first language | 403 (93.7) |
| Born in Australia | 362 (84.2) |
| Cohabitation with other adults |  |
| Live with husband or partner only | 383 (89.1) |
| Live with other family or friends including parents (husband/partner's or own) | 36 (8.4) |
| Live alone | 11 (2.6) |

\* a health care card is available to Australian residents based on low- or restricted-income tests (i.e. low income health care card), age (i.e. Commonwealth Seniors Health Card) and carer (i.e. Foster Child Health Care Card and Ex-carer Allowance Health Care Card) eligibility. These cards provide the bearer to cheaper prescription medicines and reduced or waived fees for health care (Services Australia, 2026).

**Table 2.**
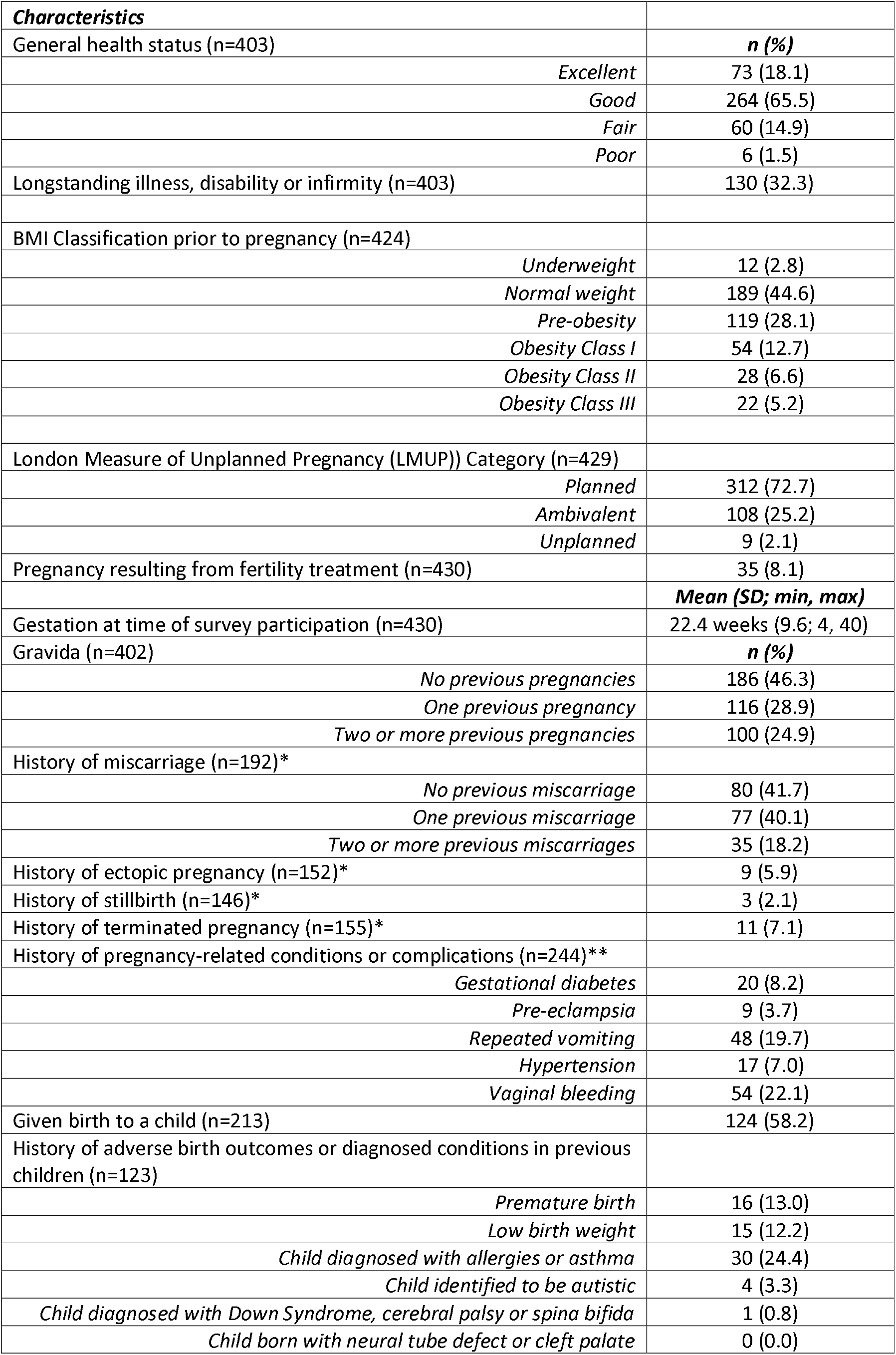
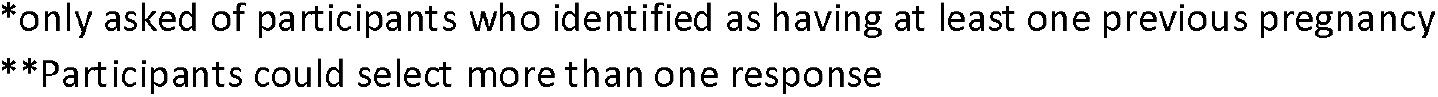
Health status, reproductive history, and pregnancy planning.

| **Characteristics** |  |
| --- | --- |
| General health status (n=403) | **n (%)** |
| *Excellent* | 73 (18.1) |
| *Good* | 264 (65.5) |
| *Fair* | 60 (14.9) |
| *Poor* | 6 (1.5) |
| Longstanding illness, disability or infirmity (n=403) | 130 (32.3) |
| BMI Classification prior to pregnancy (n=424) |  |
| *Underweight* | 12 (2.8) |
| *Normal weight* | 189 (44.6) |
| *Pre-obesity* | 119 (28.1) |
| *Obesity Class I* | 54 (12.7) |
| *Obesity Class II* | 28 (6.6) |
| *Obesity Class III* | 22 (5.2) |
| London Measure of Unplanned Pregnancy (LMUP)) Category (n=429) |  |
| *Planned* | 312 (72.7) |
| *Ambivalent* | 108 (25.2) |
| *Unplanned* | 9 (2.1) |
| Pregnancy resulting from fertility treatment (n=430) | 35 (8.1) |
|  | **Mean (SD; min, max)** |
| Gestation at time of survey participation (n=430) | 22.4 weeks (9.6; 4, 40) |
| Gravida (n=402) | **n (%)** |
| *No previous pregnancies* | 186 (46.3) |
| *One previous pregnancy* | 116 (28.9) |
| *Two or more previous pregnancies* | 100 (24.9) |
| History of miscarriage (n=192)* |  |
| *No previous miscarriage* | 80 (41.7) |
| *One previous miscarriage* | 77 (40.1) |
| *Two or more previous miscarriages* | 35 (18.2) |
| History of ectopic pregnancy (n=152)* | 9 (5.9) |
| History of stillbirth (n=146)* | 3 (2.1) |
| History of terminated pregnancy (n=155)* | 11 (7.1) |
| History of pregnancy-related conditions or complications (n=244)** |  |
| *Gestational diabetes* | 20 (8.2) |
| *Pre-eclampsia* | 9 (3.7) |
| *Repeated vomiting* | 48 (19.7) |
| *Hypertension* | 17 (7.0) |
| *Vaginal bleeding* | 54 (22.1) |
| Given birth to a child (n=213) | 124 (58.2) |
| History of adverse birth outcomes or diagnosed conditions in previous children (n=123) |  |
| *Premature birth* | 16 (13.0) |
| *Low birth weight* | 15 (12.2) |
| *Child diagnosed with allergies or asthma* | 30 (24.4) |
| *Child identified to be autistic* | 4 (3.3) |
| *Child diagnosed with Down Syndrome, cerebral palsy or spina bifida* | 1 (0.8) |
| *Child born with neural tube defect or cleft palate* | 0 (0.0) |

\*only asked of participants who identified as having at least one previous pregnancy
\*\*Participants could select more than one response

**Table 3.** Preconception health behaviours.

| Health behaviours |  |
| --- | --- |
| Use of supplements before pregnancy | **N (%)** |
| *Folate (n=390)* | 193 (49.5) |
| *Pregnancy multivitamin (n=389)* | 197 (50.6) |
| *General multivitamin (n=311)* | 49 (15.8) |
| *Vitamin D (n=322)* | 85 (26.4) |
| *Iron (n=320)* | 88 (27.5) |
| *Omega-3 fatty acids (n=309)* | 47 (15.2) |
| *Vitamin C (n=307)* | 51 (16.6) |
| *Zinc (n=298)* | 22 (7.4) |
| *Iodine (n=301)* | 32 (10.6) |
| *Calcium (n=294)* | 27 (9.2) |
| *Herbal medicine (n=299)* | 26 (8.7) |
| *Any preconception folate (n=421)\** | 281 (66.8) |
| Other preconception health behaviours |  |
| *Follow a weight loss diet (n=430)* | 97 (22.6) |
| *Get tested for a sexually transmitted infection (n=430)* | 125 (29.1) |
| *Visit the dentist (n=430)* | 202 (47.0) |
| *Check immunisation status (n=430)* | 161 (37.4) |
| *Undergo surgery to help lose weight (n=430)* | 13 (3.0) |
| Exercise behaviours |  |
| *Any exercise in the three months before conception (n=429)* | 328 (76.5) |
|  | **Mean (SD; min, max)** |
| *Hours of exercise per week (n=327)\*\** | 6.4 (5.8; 0.5, 75) |
| Smoking behaviours | **N (%)** |
| *History of having ever smoked (n=430)* | 136 (31.6) |
| *Not smoking in the three months prior to conception (n=136)\*\*\** | 84 (61.8) |
| Alcohol intake behaviours |  |
| *History of ever drinking alcohol (n=430)* | 404 (94.0) |
| *Consumed any alcohol in the three months prior to conception (n=399)\*\*\*\** | 321 (80.5) |
|  | **Mean (SD; min, max)** |
| *Total average units of alcohol consumed per week in the three months prior to conception (n=311)\*\*\*\*\** | 5.4 (5.7; 0.5, 40) |

\*includes intake of a folate-specific supplement, pregnancy multivitamin or general multivitamin
\*\* only asked of participants who reported having exercised in the three months before conception
\*\*\* only asked of participants who reported having ever smoked
\*\*\*\*only asked of participants who reported having ever drunk alcohol
\*\*\*\*\*only asked of participants who reported drinking alcohol in the three months prior to conception

### Beliefs about preconception health behaviours

The median scores for women’s beliefs regarding the importance of preconception exercise, healthy eating and alcohol avoidance varied between behaviours (see Table 4). Subjective norms for all behaviours commonly scored an average of 3 or 4 on the scale, although participants’ perceptions of their health care provider regarding these preconception health behaviours as important were higher (Median: 1).

**Table 4.** Preconception behaviour beliefs based on the Theory of Planned Behaviour. All items scored on a 7-point (0 - 7) scale in which 0 is most affirmative and 7 is least affirmative.

| CONSTRUCT |  | Regular exercise (n=430) | Eat a healthy diet (n=419) | Avoid alcohol (n=418) |
| --- | --- | --- | --- | --- |
|  |  | Median (Q1, Q3) | Median (Q1, Q3) | Median (Q1, Q3) |
| Past behaviour | I [behaviour] for at least 6 months before becoming pregnant [definitely true/definitely false] | 2 (1,5) | 3 (1, 4) | 6 (3, 7) |
| Beliefs | My doing [behaviour] during the 6 months prior to becoming pregnant is |  |  |  |
|  | [important/not important] (n=417) | 2 (1, 4) | 2 (1, 4) | 4 (1, 5) |
|  | [pleasant/unpleasant] (n=413) | 2 (1, 4) | 2 (1, 3) | 3 (1, 5) |
|  | [good/bad] (n=385) | 1 (0, 2) | 1 (0, 2) | 1 (0, 3) |
|  | If I [behaviour] for 6 months before becoming pregnant my child will be healthy [extremely likely/extremely unlikely] (n=427) | 3 (2, 4) | 3 (1, 4) | 3 (1, 4) |
| Intentions | I intend to [behaviour] for at least 6 months before becoming pregnant [extremely likely/extremely unlikely] (n=422) | 3 (1, 4) | 2 (1, 4) | 4 (1, 6) |
|  | I will make an effort to [behaviour] for at least 6 months before becoming pregnant [extremely likely/extremely unlikely] (n=411) | 2 (1, 4) | 2 (1, 3) | 3 (1, 5) |
| Control | I am confident I can [behaviour] for at least 6 months before becoming pregnant [extremely likely/extremely unlikely] (n=416) | 3 (1, 5) | 2 (1, 4) | 3 (1, 5) |
|  | [Behaviour] for 6 months before becoming pregnant is up to me [extremely likely/extremely unlikely] (n=392) | 1 (0, 2) | 1 (0, 2) | 1 (0, 1) |
| Norms | Most people like me [behaviour] for 6 months before becoming pregnant [extremely likely/extremely unlikely] (n=440) | 3 (2, 4) | 4 (3, 6) | 4 (3, 6) |
| Subjective Norms | My reproductive partner thinks it's important for me to [behaviour] for 6 months before becoming pregnant [strongly agree/strongly disagree] (n=421) | 3 (1, 4) | 2 (1, 4) | 4 (2, 5) |
|  | My older female relatives (e.g., mother, grandmother, aunt) think it's important for me to [behaviour] for 6 months before becoming pregnant [strongly agree/strongly disagree] (n=427) | 3 (2, 5) | 3 (2, 5) | 4 (2, 5.5) |
|  | My older male relatives (e.g., father, grandfather, uncle) think it's important for me to [behaviour] for 6 months before becoming pregnant [strongly agree/strongly disagree] (n=427) | 4 (2, 5) | 3 (2, 5) | 4 (3, 6) |
|  | My male friends and family of similar age think it's important for me to [behaviour] for 6 months before becoming pregnant [strongly agree/strongly disagree] (n=428) | 4 (2, 5) | 4 (2, 5) | 4 (3, 6) |
|  | My female friends and family of similar age think it's important for me to [behaviour] for 6 months before becoming pregnant [strongly agree/strongly disagree] (n=428) | 3 (2, 4) | 2 (2, 4) | 3 (2, 5) |
|  | My health care provider thinks it's important for me to [behaviour] for 6 months before becoming pregnant [strongly agree/strongly disagree] (n=412) | 1 (1, 2.5) | 1 (1, 2) | 2 (1, 3) |

### Relationships between beliefs, intentions and behaviours for preconception alcohol use, regular exercise and healthy eating

Internal reliability analyses demonstrated good to excellent consistency for most constructs, particularly beliefs (α=0.90), intentions (α=0.86), and subjective norms (α=0.94). Perceived behavioural control showed acceptable reliability (α=0.74) (see Appendix A, Table A.2).

The key path analysis and structured equation modelling outputs are reported in Table 5 and presented in Figures 1 - 3. Model fit for the alcohol avoidance structural equation modelling was good (RMSEA = 0.00, CFI = 0.98, TLI = 0.98, and SRMR = 0.03). Women’s intention to avoid alcohol in the preconception period was associated with higher levels of alcohol use behaviour (β=0.64, p<0.001). There was good evidence that combined subjective norms of health care professionals and partner were associated with higher levels of intention (β=0.31, p<0.001), as well as control (β=0.27, p<0.001) and beliefs (β=0.47, p<0.001).

**Figure 1.**
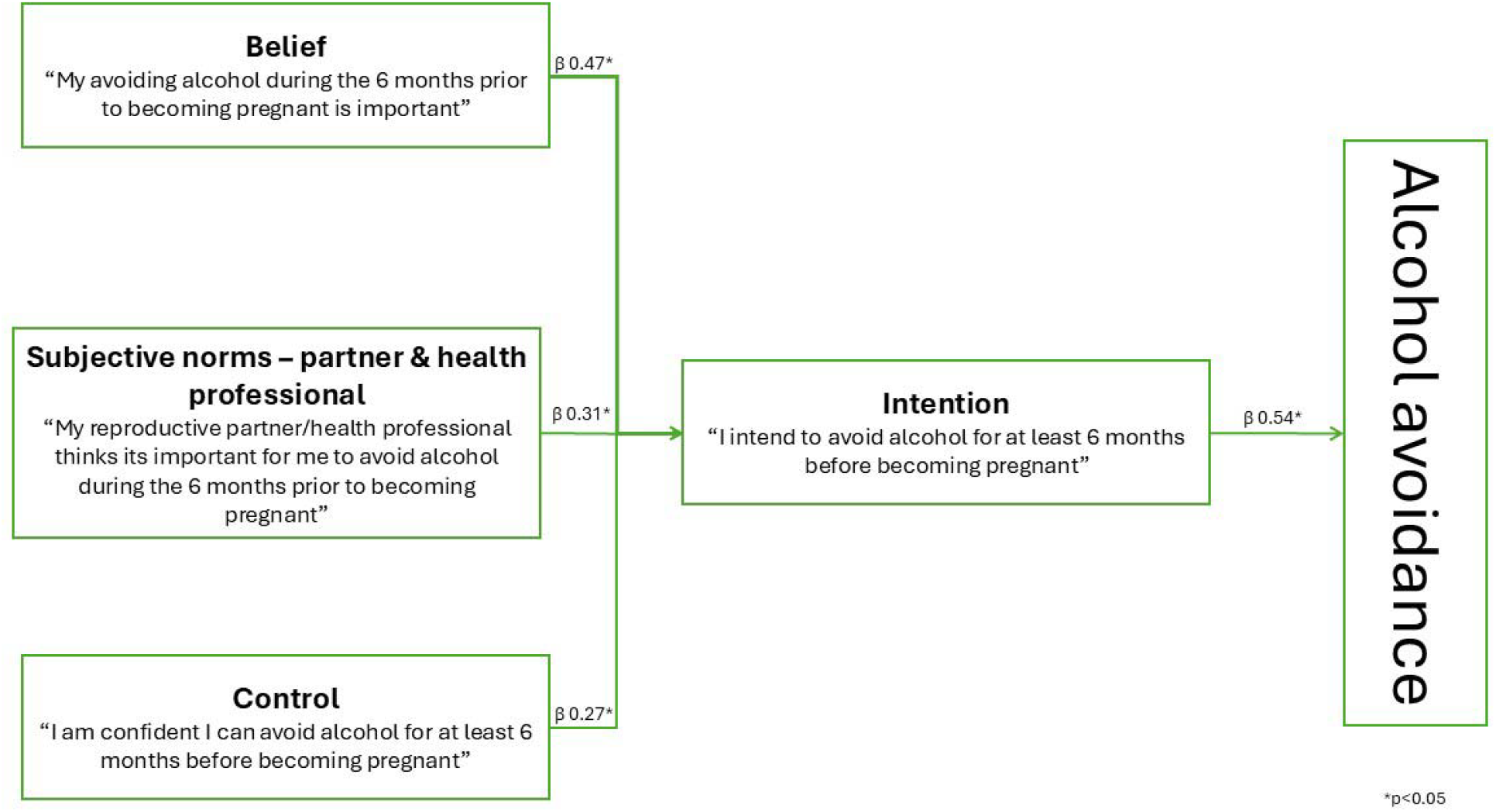
Structural equation modelling output for alcohol avoidance.

**Figure 2.**
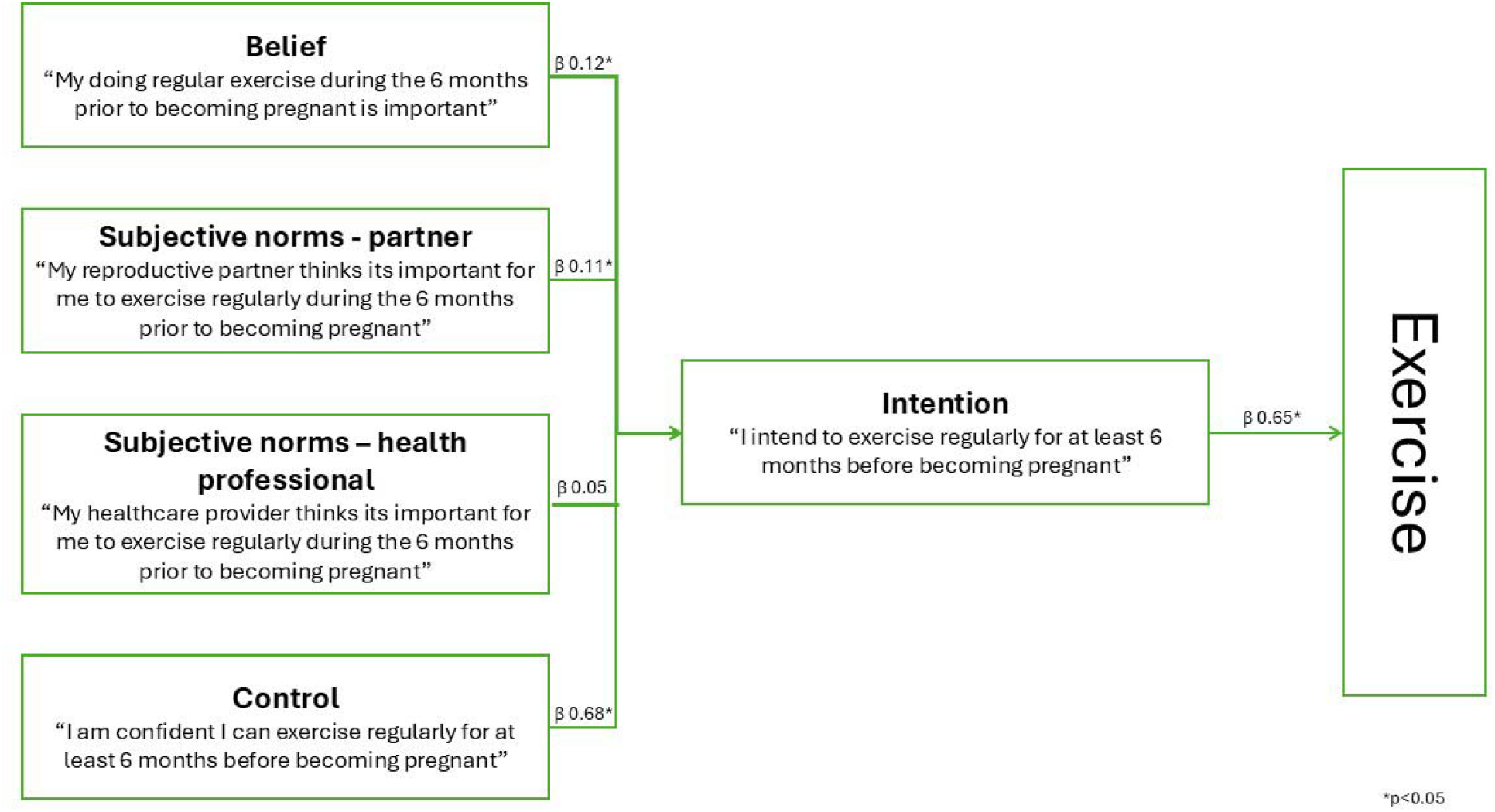
Key path analysis for regular exercise.

**Figure 3.**
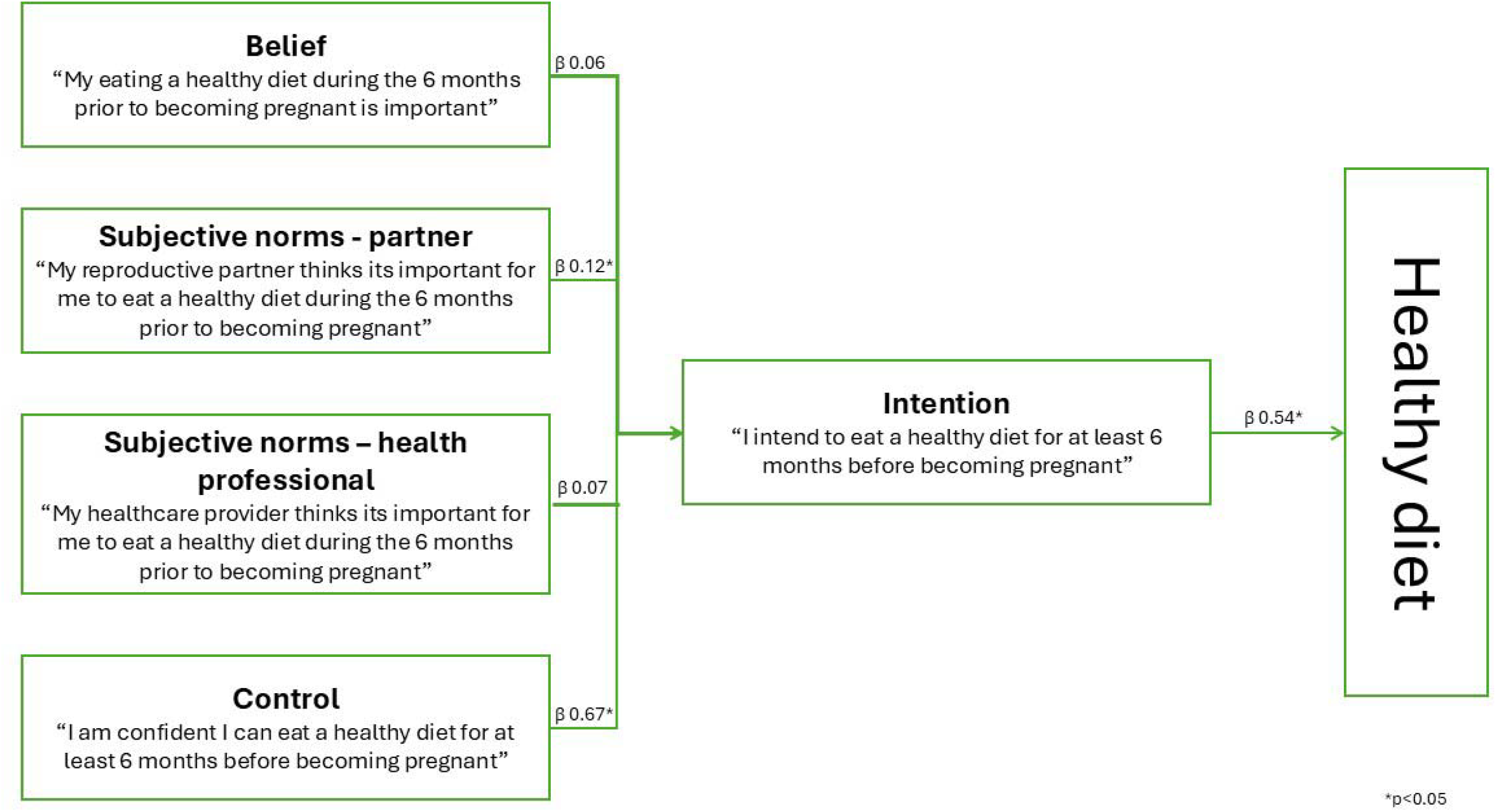
Key path analysis for eating a healthy diet.

**Table 5.** Logistic regression analysis of association between beliefs and other preconception health behaviours.

| Beliefs | Median (Q1, Q3)** | Behaviour | n (%) | Unadjusted |  | Adjusted* |  |
| --- | --- | --- | --- | --- | --- | --- | --- |
|  |  |  |  | OR (95% CI) | p | OR (95% CI) | p |
| If I <u>maintain a healthy weight</u> for 6 months before becoming pregnant my child will be healthy (n=413) | 5 (4, 6) | Maintained 'normal' BMI one month before pregnancy (n=424) | 189 (44.6) | 1.1 (0.9-1.2) | 0.09 | 1.1 (0.9-1.2) | 0.3 |
| If I <u>increase folate intake</u> for 6 months before becoming pregnant my child will be healthy (n=399) | 5 (1, 3) | Took folate supplement before pregnancy (n=421) | 281 (66.8) | 1.4 (1.3-1.6) | <0.001 | 1.4 (1.2-1.6) | <0.001 |
| If I <u>take good care of my health</u> for 6 months before becoming pregnant my child will be healthy (n=409) | 6 (5, 7) | Current self-reported health is 'excellent' or 'good' (n=403) | 337 (83.6) | 1.0 (0.8, 1.2) | 1.0 | 0.9 (0.8-1.1) | 0.6 |
| If I <u>do not smoke</u> for 6 months before becoming pregnant my child will be healthy (n=381) | 7 (6, 8) | Previous smoker did not smoke 3 months before pregnancy (n=136) | 84 (61.8) | OR 1.3 (1.1, 1.6) | 0.007 | 1.3 (1.0-1.6) | 0.03 |

\*controlled for age, relationship status, education level, financial manageability, employment status, cohabitation
\*\*1 = extremely unlikely, 7 = extremely likely

Path analysis found women’s intention with regards to preconception exercise (β=0.65, p<0.001) had a strong, positive influence on their behaviour. Their own beliefs (β=0.12, p<0.001) and control (β=0.68 p<0.001), and their perceptions of their partner’s beliefs (β=0.11, p= 0.003), also positively influenced their preconception exercise intention. There was no evidence (β=0.12, p=0.2) that subjective norms from their health professional’s beliefs influenced their intention to exercise.

Path analysis of women’s preconception diet behaviour found intention (β=0.54, p<0.001) to have a strong, positive influence on their behaviour. There was good evidence that women’s control (β=0.67, p<0.001) and subjective norms of their partner (β=0.12, p<0.001) were associated with higher levels of intention, but not the perceived subjective norms of their health care professional (β=0.07, p=0.0638), or their own beliefs (β=0.06, p=0.0793)

### Relationships between beliefs and other preconception health behaviours

Table 6 presents the regression analyses examining the association between beliefs and other preconception behaviours. The analysis found an increased likelihood that participants took a folate supplement before pregnancy if they believed it will result in their child being healthy (aOR 1.4; 95% CI 1.2, 1.6). Participants with a history of smoking were more likely to not smoke before pregnancy if they believed it would result in their child being healthy (aOR 1.3; 95% CI 1.1, 1.6). The adjusted odds ratios were not significant for beliefs associated with maintaining a healthy weight and taking good care of their health in general.

## Discussion

This analysis of the relationships between preconception health beliefs of pregnant women and their most recent preconception health behaviours identified several key findings. Firstly, the analysis identifies a strong association between participant’s intention and preconception behaviours regarding exercise, healthy eating and alcohol avoidance. Participants’ perceptions of their control regarding exercise and health diet behaviours were the strongest predictors of the associated intentions, more so than their belief in the importance of the behaviour. This is an important finding as it emphasises the critical importance of structural determinants of health in addressing preconception health behaviours as identified in previous preconception health behaviour research (Kandel et al., 2021). While it can be argued that control is an individual characteristic, such a position dismisses the external factors that impact the areas of an individual’s life within their control (Ross & Mirowsky, 2012). Research investigating factors that influence healthy eating in the general population can include food environment (e.g., food price, availability, characteristics), lived environment (e.g., time, convenience, built and natural environments), social networks (e.g., marketing and media, sociocultural acceptability) as well as more individual factors (e.g., knowledge and skills, psychology, physiology, habits) (Zorbas et al., 2018). While there are some differences in motivations for health behaviour change in pregnancy, whereby a woman may navigate potentially conflicting desires for their own body and for the health of their child, external influences (e.g. practical, environmental, social) remain (Rockliffe et al., 2021).

Another key finding is that perceived control was not influential in alcohol avoidance behaviour in the study population. The reason for this preconception health behaviour dynamic warrants closer examination but may be affected by strong social and cultural factors within the Australian community, particularly with regards to the normalisation of drinking and perceptions that alcohol intake is an expected behaviour in social gatherings (Palmer et al., 2024). Poor understanding of drinking guidelines also influence drinking behaviour in Australian adults (Palmer et al., 2024) and Australian women of reproductive age consider alcohol avoidance in the preconception period to be less relevant and important than during pregnancy (Musgrave et al., 2023; Smith et al., 2022). With this in mind, there is a clear need to not only provide women with the information they require to accurately understand the implications of their preconception health behaviours (Caut et al., 2022) but also, in parallel, to develop policy changes and funding of health and social services which make changing such health behaviours more accessible for women (Steel et al., 2025).

It is also interesting to note that, according to this study’s findings, while women’s perceptions of their reproductive partner’s beliefs about their preconception health exercise, diet and alcohol behaviours was associated with the woman’s intention to change that behaviour, the beliefs of their health professional relating to these behaviours were not. This is particularly interesting given the median scores for reproductive partners’ beliefs were lower than health professionals’ beliefs for all three behaviours. This finding presents challenges to existing attempts to implement preconception health interventions, which usually centre on clinical contexts. With this in mind, women’s preconception health behaviour change may require more active involvement from and education of their partner. However, men experience specific barriers to engaging with preconception care ranging from health professionals lacking understanding and confidence discussing preconception care with men (Carter et al., 2025; Hogg et al., 2019) through to men feeling that they are unwelcome in the health care encounter associated with their partner’s pregnancy which may extend to the preconception period (Steen et al., 2012). Yet there is an urgent need to overcome these challenges as partners not only have a role in supporting women to make positive health behaviour change (Rockliffe et al., 2021), but men’s preconception health behaviours also have a direct physiological and epigenetic effect on the health of the child at birth and later life (Carter et al., 2023; Huang et al., 2026). While men’s attitudes and behaviours with regards to preconception health behaviours have not received the same research attention as women’s (Carter et al., 2023, 2025), there is growing evidence that steps can be taken to improve men’s engagement in this critical life stage (Agustina et al., 2024). Any attempts to address this must, however, also consider the documented concerns among women with regards to women’ rights to bodily autonomy, particularly within the context of pregnancy and birth (Rockliffe et al., 2021). Health promotion and public health practitioners are, therefore, faced with the difficulty of supporting women’s intention to improve their preconception health behaviour, whilst not weaponising the importance of preconception behaviour change. One possible solution is to equally educate men about the importance of their own preconception health behaviour (Carter et al., 2023; Steel & Carter, 2021). Overall, our findings suggest future preconception health interventions need to be couples-focused to achieve the best possible outcomes.

The logistic regression analysis of other preconception health behaviours found folate supplement use and smoking cessation to be likely to change in alignment with preconception health beliefs whereas maintaining a healthy weight or taking good care of health was not. This difference may be because folate intake and smoking cessation have received attention in health promotion messaging for preconception and pregnancy in recent years (Barker et al., 2018), however it is also worth noting that not all participants for which these behaviours were relevant actually reported those behaviours. While it is possible that messages regarding folate and smoking cessation may not have reached all population groups, it is also possible that factors similar to those identified for alcohol, diet and exercise (e.g. perceived control or partner attitudes) have a role in women’s decision-making for these two behaviours (Kandel et al., 2021). In contrast, having a normal weight or good health during preconception were not associated with women’s beliefs about the importance of these two factors. This variation may be due to body weight and good health being health ‘states’ rather than health ‘behaviours’. Although maintaining these states may require active behaviours, they also call on women to make behaviour change for a longer period of time and require a greater time commitment towards preconception health improvement compared to taking a vitamin supplement or quitting smoking (Stephenson et al., 2018). It also may be that there is additional complexity – biological, psychological, and social – to women attaining these health states (Harrison et al., 2017; Williams et al., 2024) which may be further exacerbated, in the case of body weight, by stigma and blame affecting the level of support provided to individuals with overweight and obesity (Hill & Rodriguez, 2020; Kirk et al., 2014). However, even after controlling for a range of sociodemographic factors, the relationship remained. Given the need for tailored, specialised preconception health care for individuals with compromised health or diagnosed health conditions, and equally for those with obesity, the factors that drive women’s ability to address these health states requires closer attention and the time required to achieving optimal states prior to conception warrants further research.

### Implications for Practice and/or Policy

This analysis identifies two key areas that have direct and critical implications for practice and policy. Firstly, the finding that a woman’s perceptions of their health professional’s beliefs about the importance of the woman’s preconception health behaviours is a novel and important finding. A considerable amount of research effort has been focused on strengthening health professional education and skill with regards to delivering preconception care. While this work is undoubtedly important and necessary to ensure women have access to safe and effective care (Caut et al., 2025a), the role of this care in facilitating a woman’s behaviour change may need to be reconsidered. In parallel, the finding that a woman’s perception of their partner’s beliefs about the importance of their behaviour change points both clinicians and policymakers to the critical need to include partners in the preconception care journey. This need has received growing attention in recent years in light of the impacts of non-pregnant partner preconception behaviour on birth and offspring outcomes (Carter et al., 2023), yet our analysis highlights the importance of the non-pregnant partner in a couple irrespective of their role in conception. This finding is important for clinicians as current evidence suggests they rely on women relaying information to their partner, rather than directly involving the partner in the care journey (Caut et al., 2025a, 2025b). Policymakers must also be aware of this dynamic and take a broader approach to addressing preconception health behaviour change in the community. This includes implementing public-facing health promotion campaigns to educate reproductive couples about their shared role in pregnancy preparation, and addressing ongoing structural factors that limit individual’s control over health behaviour change such as access to health care, housing, financial security and other critical drivers underpinning locus of control (Steel et al., 2025).

### Limitations

The results of this analysis must be interpreted within the context of their limitations. The participants’ responses are self-reported and are exposed to potential responder bias. Also, the behaviours included in the modelling are past behaviours, which were used as a proxy for future behaviour based on current knowledge and beliefs, but these predictors may have altered since the time of the past behaviours. It is also notable that it was only possible to conduct structural equation modelling for alcohol avoidance, and as such different analyses was applied to other behaviours. Further, the model did not permit analysis of other possible social norms, and therefore the TPB model was not able to be tested in full. There are also numerous behavioural theories which may be applied to this research question and the appropriateness of TPB has been queried for evaluating pregnancy intention (Stewart & Hall, 2024). However, given this analysis was aimed at investigating health behaviours for which the TPB has a longstanding history of application, the results should still be considered valid.

### Conclusions

This study reinforces the central role of intention in shaping preconception health behaviours, while highlighting perceived control and partner influences as key determinants of that intention. These findings underscore the importance of addressing structural and social contexts, beyond individual knowledge, to enable meaningful behaviour change. The limited influence of health professionals, alongside the stronger role of partners, suggests a need to reorient interventions toward more inclusive, couple-focused approaches. Differences observed across behaviours further indicate that not all preconception practices are influenced in the same way, particularly for more complex health states such as body weight. Future research should explore these distinctions and the broader barriers women face, while policy and practice should prioritise accessible, supportive environments that facilitate sustained behaviour change before conception.

## Data Availability

All data produced in the present study are available upon reasonable request to the authors

## Appendix A

**Table A.1.** Differences in participant demographics among those included in the analysis and the full sample.

| Characteristics | All respondents<br>(n=617)<br>n (%) | Included in analysis<br>(n=430)<br>n (%) | p value<br>(Cramer's V) |
| --- | --- | --- | --- |
| Relationship status (n=430) |  |  |  |
| Married | 327 (53.0) | 240 (55.8) | 0.07 |
| De facto | 257 (41.7) | 168 (39.1) |  |
| Other | 33 (5.4) | 22 (5.1) |  |
| Highest level of education (n=430) |  |  |  |
| Year 10 or less | 22 (3.6) | 11 (2.6) | 0.003<br>(V=0.1491) |
| Year 12 or equivalent | 33 (7.3) | 31 (7.2) |  |
| Apprenticeship, certificate or diploma | 116 (25.7) | 115 (26.7) |  |
| University qualification | 292 (64.6) | 273 (63.5) |  |
| Financial manageability (n=430) |  |  |  |
| It is impossible/It is difficult all the time | 37 (6.0) | 25 (5.8) | 0.9 |
| It is difficult some of the time | 97 (21.5) | 95 (22.1) |  |
| It is not too bad | 211 (46.7) | 198 (46.1) |  |
| It is easy | 118 (26.1) | 112 (26.1) |  |
| Employment status (n=430) |  |  |  |
| Full time work (35 hours or more per week) | 320 (51.9) | 227 (52.8) | 0.08 |
| Part time work (less than 35 hours per week) | 151 (24.5) | 108 (25.1) |  |
| Casual or temporary work (irregular hours) | 58 (9.4) | 37 (8.6) |  |
| Currently looking for work | 16 (2.6) | 8 (1.9) |  |
| Not currently in the paid workforce and not looking for work | 72 (11.7) | 50 (11.6) |  |
| Private health insurance cover (n=430) |  |  |  |
| No | 267 (43.3) | 179 (41.6) | 0.07 |
| Yes, but no obstetrics cover | 186 (30.2) | 138 (32.1) |  |
| Yes, with obstetrics cover | 164 (26.6) | 113 (26.3) |  |
| Health Care Card* (n=430) | 91 (20.1) | 87 (20.2) | 0.003<br>(V=0.1194) |
| Aboriginal or Torres Strait Islander status |  |  |  |
| No | 581 (94.2) | 401 (93.3) | 0.6 |
| Yes | 30 (4.9) | 24 (5.6) |  |
| I would rather not say | 6 (1.0) | 5 (1.2) |  |
| English as first language | 574 (93.0) | 403 (93.7) | 0.2 |

\*Cramer's V, strength of association: > 0.25 = very strong, > 0.15 = strong, > 0.10 = moderate, > 0.05 = weak association, > 0 = weak.

**Table A.2.** Internal Reliability analysis.

| Construct | Items | Mean | SD | $\alpha$ | |
| --- | --- | --- | --- | --- | --- |
| Past Behaviours | 3 | 3.48 | 1.66 | 0.47 |  |
| Beliefs | 16 | 2.50 | 1.21 | 0.90 |  |
| Intentions | 6 | 2.83 | 1.69 | 0.86 |  |
| Control | 6 | 2.08 | 1.35 | 0.74 |  |
| Norms | 3 | 3.54 | 1.56 | 0.78 |  |
| Subjective Norms | 18 | 3.15 | 1.38 | 0.94 |  |
| **Beliefs** |  |  |  |  |  |
| *Exercise regularly* | 4 | 2.40 | 1.44 | 0.73 |  |
| *Healthy diet* | 4 | 2.29 | 1.33 | 0.76 |  |
| *Avoiding alcohol* | 4 | 2.97 | 1.71 | 0.79 |  |
| **Subjective norms** |  |  |  |  |  |
| *Exercise regularly* | 6 | 3.06 | 1.44 | 0.88 |  |
| *Healthy diet* | 6 | 2.80 | 1.43 | 0.88 |  |
| *Avoiding alcohol* | 6 | 3.54 | 1.79 | 0.91 |  |
| **Intentions** | **Items** | **Mean** | **SD** | ***r*** | ***p*** |
| *Exercise regularly* | 2 | 2.71 | 1.98 | 0.80 | <0.001 |
| *Healthy diet* | 2 | 2.34 | 1.79 | 0.84 | <0.001 |
| *Avoiding alcohol* | 2 | 3.37 | 2.37 | 0.83 | <0.001 |
| **Control** |  |  |  |  |  |
| *Exercise regularly* | 2 | 2.18 | 1.67 | 0.38 | <0.001 |
| *Healthy diet* | 2 | 1.93 | 1.45 | 0.38 | <0.001 |
| *Avoiding alcohol* | 2 | 2.07 | 1.70 | 0.28 | <0.001 |

*r* = Pearson's correlation coefficient
Strong internal reliability = $\alpha > 0.70$

**Table A.3.** Structural equation modeling and key path analysis using the Theory of Planned Behaviour to identify predictors for preconception exercise, diet and alcohol behaviours.

| Outcome | Covariate | Standardised estimate | Lower CL | Upper CL | p-value |
| --- | --- | --- | --- | --- | --- |
| **Alcohol use (Structural equation modeling)** |  |  |  |  |  |
| Behaviour | Intention | 0.64 | 0.57 | 0.70 | <0.001 |
| Intention | Belief | 0.47 | 0.34 | 0.61 | <0.001 |
| Intention | Subjective Norms | 0.31 | 0.21 | 0.40 | <0.001 |
| Intention | Control | 0.27 | 0.17 | 0.37 | <0.001 |
| **Regular exercise (Key path analysis)** |  |  |  |  |  |
| Behaviour | Intention | 0.65 | 0.60 | 0.71 | <0.001 |
| Intention | Belief | 0.12 | 0.05 | 0.18 | <0.001 |
| Intention | Subjective Norms - partner | 0.11 | 0.04 | 0.18 | 0.0032 |
| Intention | Subjective Norms - HCP | 0.05 | -0.02 | 0.12 | 0.1891 |
| Intention | Control | 0.68 | 0.62 | 0.73 | <0.001 |
| **Healthy eating (Key path analysis)** |  |  |  |  |  |
| Behaviour | Intention | 0.54 | 0.48 | 0.61 | <0.001 |
| Intention | Belief | 0.06 | -0.01 | 0.13 | 0.0793 |
| Intention | Subjective Norms - partner | 0.12 | 0.05 | 0.20 | <0.001 |
| Intention | Subjective Norms - HCP | 0.07 | 0.00 | 0.14 | 0.0638 |
| Intention | Control | 0.67 | 0.61 | 0.73 | <0.001 |

